# Extensive weight loss can reduce immune age by altering IgG N-glycosylation

**DOI:** 10.1101/2020.04.24.20077867

**Authors:** Valentina L Greto, Ana Cvetko, Tamara Štambuk, Niall J Dempster, Domagoj Kifer, Helena Deriš, Ana Cindrić, Frano Vučković, Mario Falchi, Richard S Gillies, Jeremy W Tomlinson, Olga Gornik, Bruno Sgromo, Tim D Spector, Cristina Menni, Alessandra Geremia, Carolina V Arancibia-Cárcamo, Gordan Lauc

## Abstract

**Background:** Obesity is a major global health problem, and is associated with increased cardiometabolic morbidity and mortality. Protein glycosylation is a frequent postranslational modification, highly responsive to numerous pathophysiological conditions and ageing. The prospect of biological age reduction, by reverting glycosylation changes through metabolic intervention, opens many possibilities. We have investigated whether weight loss interventions affect inflammation- and ageing-associated IgG glycosylation changes, in a longitudinal cohort of bariatric surgery patients. To support potential findings, BMI-related glycosylation changes were monitored in a longitudinal twins cohort.

**Methods:** IgG N-glycans were chromatographically profiled in 37 obese patients, subjected to low-calorie diet, followed by bariatric surgery, across multiple timepoints. Similarly, plasma-derived IgG N-glycan traits were longitudinally monitored in 1,680 participants from the TwinsUK cohort.

**Results:** Low-calorie diet induced a marked decrease in the levels of IgG N-glycans with bisecting GlcNAc, whose higher levels are usually associated with ageing and inflammatory conditions. Bariatric surgery resulted in extensive alterations of the IgG glycome that accompanied progressive weight loss during one-year follow-up. We observed a significant increase in digalactosylated and sialylated glycans, and a substantial decrease in agalactosylated and core fucosylated IgG glycans. In general, this IgG glycan profile is associated with a younger biological age and reflects an enhanced anti-inflammatory IgG potential. Loss of BMI over a 20 year period in the TwinsUK cohort validated a weight loss-associated agalactosylation decrease and an increase in digalactosylation.

**Conclusions:** Altogether, these findings highlight that weight loss substantially affects IgG N-glycosylation, resulting in reduced biological and immune age.

**GRAPHICAL ABSTRACT:** 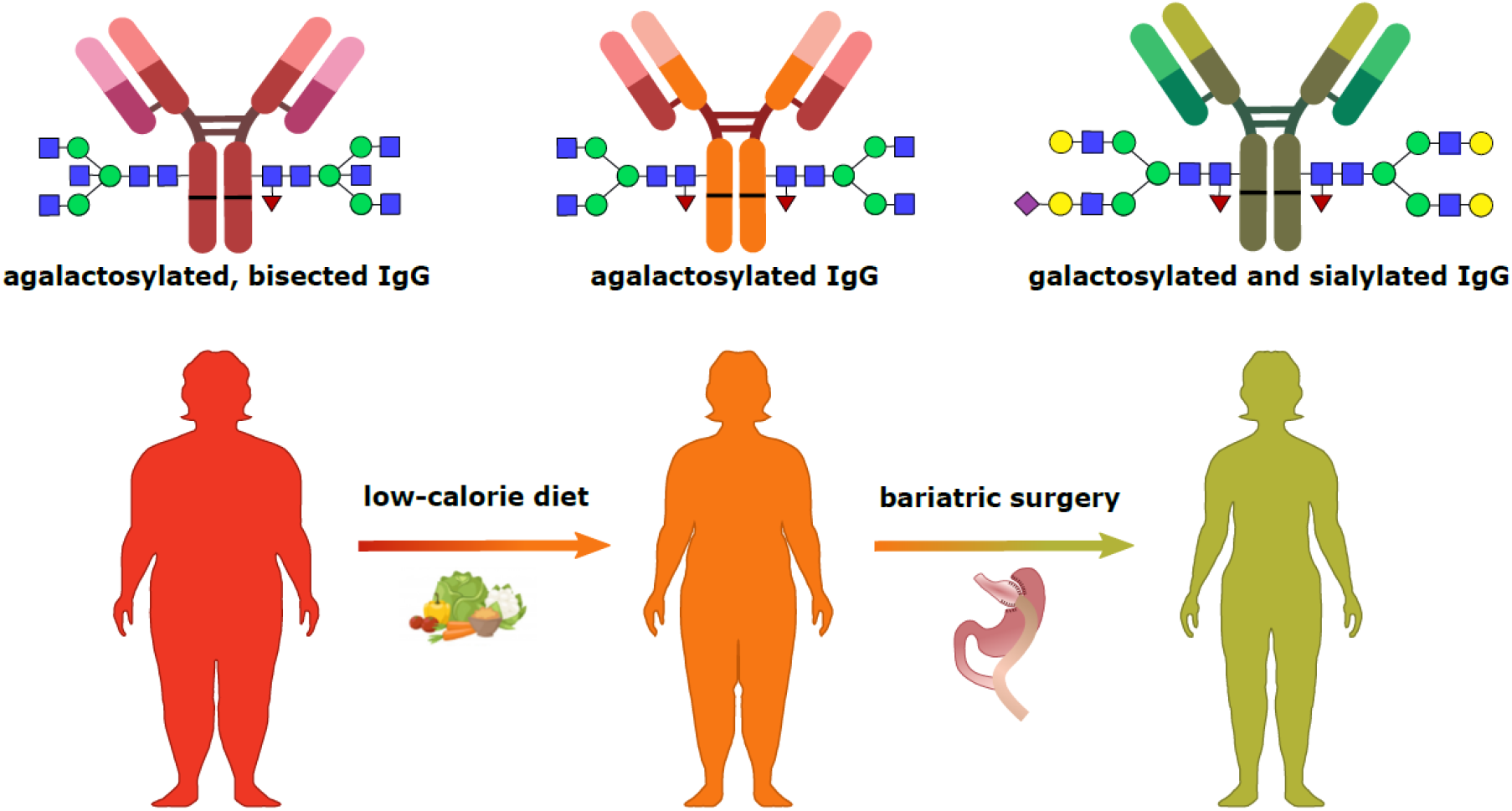

**HIGHLIGHTS:**

- Obesity is associated to inflammation-related agalactosylated and bisected IgG glycoforms
- IgG galactosylation and sialylation increase after bariatric surgery-induced weight loss
- Progressive decrease of BMI is associated to increased IgG galactosylation, implying a reduction of biological age

## INTRODUCTION

The global prevalence of obesity has risen dramatically in the past decades, and it is now considered a pandemic [1]. According to the World Health Organization, over 650 million individuals are obese, accounting for 13% of the world’s adult population. Obesity confers a risk for metabolic syndrome, contributing to type 2 diabetes and cardiovascular disease (CVD) development [2]. Metabolic syndrome is linked to a chronic systemic low-grade inflammation, which contributes to the aging of the immune system denoted as inflammaging [3,4]. Obesity-related inflammaging results in impaired innate and adaptive immune function, and is characterized by high serum levels of IL-6, TNF-α and CRP [5]. Altered protein N-glycosylation is one of the hallmarks of inflammaging [4,6]. The human circulating N-glycome represents the entire set of glycans that are covalently attached to plasma proteins through a nitrogen on an asparagine residue. N-glycans are essential for life and are involved in many physiological processes [7], including signal transduction, protein trafficking and folding, receptor regulation and cell adhesion. Glycosylation has a fundamental role in the innate and adaptive immune responses, accentuated by the fact that all five classes of immunoglobulins (Ig) bear N-glycans. In this regard, IgG is probably the most investigated glycoprotein, whose effector functions are controlled by its Fc-bound glycans [8].

Inter-individual differences in the pace of biological ageing is an intriguing concept which may contribute to why some people stay healthy until their late chronological age, while others age faster and have shorter life expectancy. Progressive age-related changes of IgG glycosylation have been extensively studied [7,9,10] and the GlycanAge model has been proposed to express the difference between chronological and IgG glycome ageing [11]. The age of the IgG glycome might be estimated through the levels of agalactosylated species, which increase with ageing and are associated with enhanced immune activation [12]. The opposite applies for digalactosylated IgG glycoforms, which are usually related to a younger age. Besides age-related changes, specific IgG glycosylation patterns have been already associated with CVD risk score and subclinical atherosclerosis in two large independent UK cohorts [13]. Moreover, a prospective follow-up of the EPIC-Potsdam cohort confirmed that changes in plasma N-glycome composition are predictive of future CVD events, with comparable predictive power to the American Heart Association (AHA) score in men and even better predictive power in women [14]. The link between a proinflammatory IgG N-glycome and hypertension has also been extensively studied [15-17], and similar IgG glycosylation patterns were associated with increased body mass index (BMI) and measures of central adiposity [18,19].

Studies in mouse models further corroborate the importance of differential IgG glycoforms in CVD pathogenesis. It has been shown that hyposialylated IgG (corresponding to an old-like IgG glycome) can induce obesity-related hypertension and insulin resistance in B-cell-deficient mice, through activation of the endothelial FcγRIIB [20,21]. These findings indicate that the IgG N-glycome could represent more than a biomarker of inflammation and aging, since distinctive IgG glycoforms act as effector molecules in certain pathologies. Furthermore, supplementation with *N*-acetylmannosamine (ManNAc), a precursor of sialic acid, protects obese mice from hypertension and insulin resistance induction by reverting an IgG N-glycome associated with old age into an IgG N-glycome associated with young age [21,22]. However, studies exploring the possibilities of converting an old-like into a young-like IgG glycome by metabolic intervention in humans are limited. Of note, only one small study indicated that high-intensity interval training can rejuvenate the IgG N-glycome [23].

Bariatric surgery is very effective for the treatment of severe obesity [24]. The resulting weight loss impacts energy balance and metabolism, contributing to the increased insulin response, improved glycaemic control and reduction of total body fat, leading to decreased CVD risk and mortality [25].

In this study we aimed to determine whether weight loss affects glycan markers related to inflammation and ageing, in a longitudinally-monitored cohort of obese individuals undergoing low-calorie diet and then bariatric surgical interventions. We also investigated BMI-related glycosylation changes in the longitudinal TwinsUK cohort, the largest cohort of adult twins with the most detailed clinical database in the world.

## METHODS

### Study populations

#### Bariatric cohort

The cohort included 37 participants, recruited at Oxford University Hospitals to the Gastrointestinal Illnesses study (Ref: 16/YH/0247). All patients were characterised by metabolic status and medical history. Bariatric patients were considered eligible in accordance to National Institute for Health and Care Excellence (NICE) and local guidelines.

Patients with a history of alcoholism and/or ongoing anticoagulant treatment were excluded. Patients were also excluded in case of pregnancy, active substance abuse or uncontrolled psychiatric condition including eating disorders. Participants were sampled at baseline and subjected to 3-week low calorie carbohydrate-restricted diet (900 cal, maximum 100 g of carbohydrates per day), followed by bariatric surgery. The sequential follow-up timepoints included the day of the surgery (baseline), at 20% of weight loss after 6.54 ± 3.4 months (mean ± IQR) and 12.47 ± 6.55 months post-op. Characteristics of the bariatric cohort are shown in Table 1.

**Table 1.**
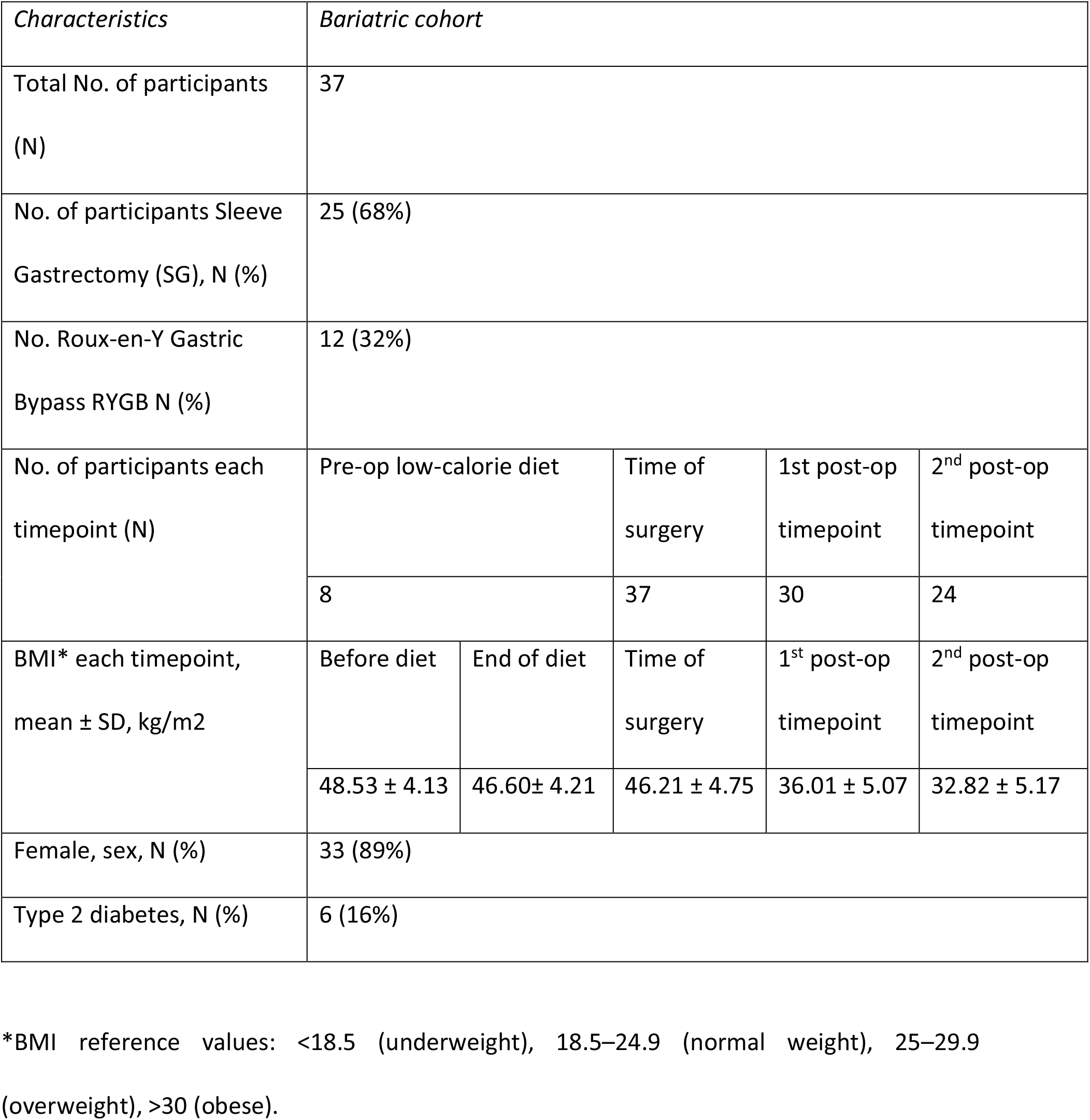
Demographic characteristics of the bariatric cohort.

#### TwinsUK cohort

We have analysed a total of 6,032 plasma samples from 2,146 participants of the TwinsUK study, collected at multiple timepoints over a 20 year-period [26]. These included 1,865 individuals sampled at 3 timepoints, 156 individuals sampled at 2 timepoints and 125 individuals sampled only once. Following the plasma N-glycome analysis, glycan data underwent quality control (see *Statistical analysis* section), which decreased the dataset to 5,889 samples (measurements). Out of these 5,889 measurements, we have proceeded with statistical analysis on a subset of 3,742 samples (measurements) which had information on BMI available. Description of the TwinsUK cohort is provided in Table 2.

### Ethical statement

Ethical approval for the study was obtained by the National Research Ethics Committees of the UK National Health Service (NHS) under the reference number 16/YH/0247. All individuals participating in this study gave written informed consent. The TwinsUK study was approved by NRES Committee London–Westminster, and all twins provided informed written consent.

### N-glycome analysis

#### Isolation of IgG from human plasma

IgG was isolated from plasma samples by affinity chromatography as described previously [27]. In brief, IgG was isolated in a high-throughput manner, using 96-well protein G monolithic plates (BIA Separations, Slovenia), starting from 100 μl of plasma. Plasma was diluted 7x with phosphate buffered saline (PBS; Merck, Germany) and applied to the protein G plate. IgG was eluted with 1 ml of 0.1 M formic acid (Merck, Germany) and immediately neutralized with 1 M ammonium bicarbonate (Acros Organics, USA).

#### N-glycan release from IgG and total plasma proteins

Isolated IgG samples were dried in a vacuum centrifuge. After drying, IgG was denatured with the addition of 30 μl of 1.33% SDS (w/v) (Invitrogen, USA) and by incubation at 65 °C for 10 min. Plasma samples (10 μl) were denatured with the addition of 20 μl of 2% (w/v) SDS (Invitrogen, USA) and by incubation at 65 °C for 10 min. From this point on, the procedure was identical for both IgG and plasma samples. After denaturation, 10 μl of 4% (v/v) Igepal-CA630 (Sigma Aldrich, USA) was added to the samples, and the mixture was shaken 15 min on a plate shaker (GFL, Germany). N-glycans were released with the addition of 1.2 U of PNGase F (Promega, USA) and overnight incubation at 37 °C.

#### Fluorescent labelling and HILIC SPE clean-up of released N glycans

The released N-glycans were labelled with 2-aminobenzamide (2-AB). The labelling mixture consisted of 2-AB (19.2 mg/ml; Sigma Aldrich, USA) and 2-picoline borane (44.8 mg/ml; Sigma Aldrich, USA) in dimethyl sulfoxide (Sigma Aldrich, USA) and glacial acetic acid (Merck, Germany) mixture (70:30 v/v). To each sample 25 μl of labelling mixture was added, followed by 2 h incubation at 65 °C. Free label and reducing agent were removed from the samples using hydrophilic interaction liquid chromatography solid-phase extraction (HILIC-SPE). After incubation samples were brought to 96% of acetonitrile (ACN) by adding 700 μl of ACN (J.T. Baker, USA) and applied to each well of a 0.2 μm GHP filter plate (Pall Corporation, USA). Solvent was removed by application of vacuum using a vacuum manifold (Millipore Corporation, USA). All wells were prewashed with 70% ethanol (Sigma-Aldrich, St. Louis, MO, USA) and water, followed by equilibration with 96% ACN. Loaded samples were subsequently washed 5x with 96% ACN. N-glycans were eluted with water and stored at – 20 °C until usage.

#### Hydrophilic interaction liquid chromatography of N-glycans

Fluorescently labelled N-glycans were separated by hydrophilic interaction liquid chromatography (HILIC) on Acquity UPLC H-Class instrument (Waters, USA) consisting of a quaternary solvent manager, sample manager and a fluorescence detector, set with excitation and emission wavelengths of 250 and 428 nm, respectively. The instrument was under the control of Empower 3 software, build 3471 (Waters, Milford, USA). Labelled N-glycans were separated on a Waters BEH Glycan chromatography column, with 100 mM ammonium formate, pH 4.4, as solvent A and ACN as solvent B. In the case of IgG N-glycans, separation method used linear gradient of 75–62% acetonitrile at flow rate of 0.4 ml/min in a 27-min analytical run. For plasma protein N-glycans separation method used linear gradient of 70–53% acetonitrile at flow rate of 0.561 ml/min in a 25-min analytical run. The system was calibrated using an external standard of hydrolysed and 2-AB labelled glucose oligomers from which the retention times for the individual glycans were converted to glucose units (GU). Data processing was performed using an automatic processing method with a traditional integration algorithm after which each chromatogram was manually corrected to maintain the same intervals of integration for all the samples. The chromatograms were all separated in the same manner into 24 peaks (GP1–GP24) for IgG N-glycans and 39 peaks (GP1–GP39) for plasma protein N-glycans and are depicted in Supplementary Figure 1 and Supplementary Figure 2, respectively. Detailed description of glycan structures corresponding to each glycan peak is presented in Supplementary Table 1. Glycan peaks were analysed based on their elution positions and measured in glucose units, then compared to the reference values in the “GlycoStore” database (available at: https://glycostore.org/) for structure assignment. The amount of glycans in each peak was expressed as a percentage of the total integrated area. For IgG glycans, in addition to 24 directly measured glycan traits, 8 derived traits were calculated (Supplementary Table 2). In the case of TwinsUK cohort, IgG N-glycan traits were calculated from plasma protein glycan profiles, based on known elution positions of predominat IgG glycan structures (Supplementary Table 3). In general, derived glycan traits average particular glycosylation features, such as galactosylation, fucosylation, bisecting GlcNAc, and sialylation.

## Statistical analysis

### Bariatric cohort

In order to remove experimental variation from the measurements, normalization and batch correction were performed on the UPLC glycan data. To make measurements across samples comparable, normalization by total area was performed. Prior to batch correction, normalized glycan measurements were log-transformed due to right-skewness of their distributions and the multiplicative nature of batch effects. Batch correction was performed on log-transformed measurements using the ComBat method (R package sva [28], where the technical source of variation (which sample was analysed on which plate) was modelled as batch covariate. To correct measurements for experimental noise, estimated batch effects were subtracted from log-transformed measurements.

Longitudinal analysis of patient samples through their observation period was performed by implementing a linear mixed effects model, where time was modelled as fixed effect, while the individual ID was modelled as random effect. Prior to the analyses, glycan variables were all transformed to standard Normal distribution by inverse transformation of ranks to Normality (R package “GenABEL”, function rntransform). Using rank transformed variables makes estimated effects of different glycans comparable, as these will have the same standardized variance. False discovery rate (FDR) was controlled by the Benjamini-Hochberg procedure at the specified level of 0.05. Data was analysed and visualized using R programming language (version 3.5.2)[29].

### TwinsUK cohort

Normalization of peak intensities to the total chromatogram area was performed for each measured sample separately. Calculated proportions were then batch corrected using ComBat method (R package sva)[28]. After the batch correction the first 11 peaks, which predominantly originate from IgG [30], were used to calculate 6 derived glycan traits – agalactosylation (G0), monogalactosylation (G1), digalactosylation (G2), bisecting GIcNAc (B), core fucosylation (CF) and high mannose structures (HM). Mixed models were fitted to estimate the effect of BMI change on IgG N-glycome (R package *lme4)*[31]. Directly measured or derived glycan trait was used as a dependent variable in the mixed model. To differentiate between BMI change and the absolute BMI value, the variable was separated to BMI_baseline_ and BMI_difference_ (calculated according to the following equation: *BMI_difference_ = BMI_follow up age_ − BMI_baseline age_*), and both were used in the model as a fixed effect. Since IgG N-glycome is affected by aging, age was included both as a fixed effect and a random slope. Finally, to meet the independency criteria, family ID and individual ID (nested within family) were included in the model as a random intercept. Due to multiple model fitting (for 11 directly measured and 6 derived glycan traits) false discovery rate was controlled using Benjamini-Hochberg method. All statistical analyses were performed using R programming language (version 3.6.3)[29].

## RESULTS

### Impact of pre-surgical low-calorie diet on IgG glycosylation

We chromatographically profiled the IgG N-glycome in a cohort of bariatric surgery-candidate patients before and after the pre-operative diet. By employing a linear mixed model, we observed significant change in only one out of eight examined IgG derived glycan traits. Namely, the levels of bisecting GlcNAc (B) were substantially decreased after the low-calorie diet intervention (Table 3), indicating a decreased proinflammatory potential of the circulating IgG. The other IgG glycosylation features did not exhibit significant alterations, possibly due to rather short follow-up period (3 weeks) and limited number of participants (n=8) (Table 3).Graphical representation of the longitudinal alterations in IgG N-glycome after low-calorie diet are depicted in Supplementary Figure 3.

**Table 2.**
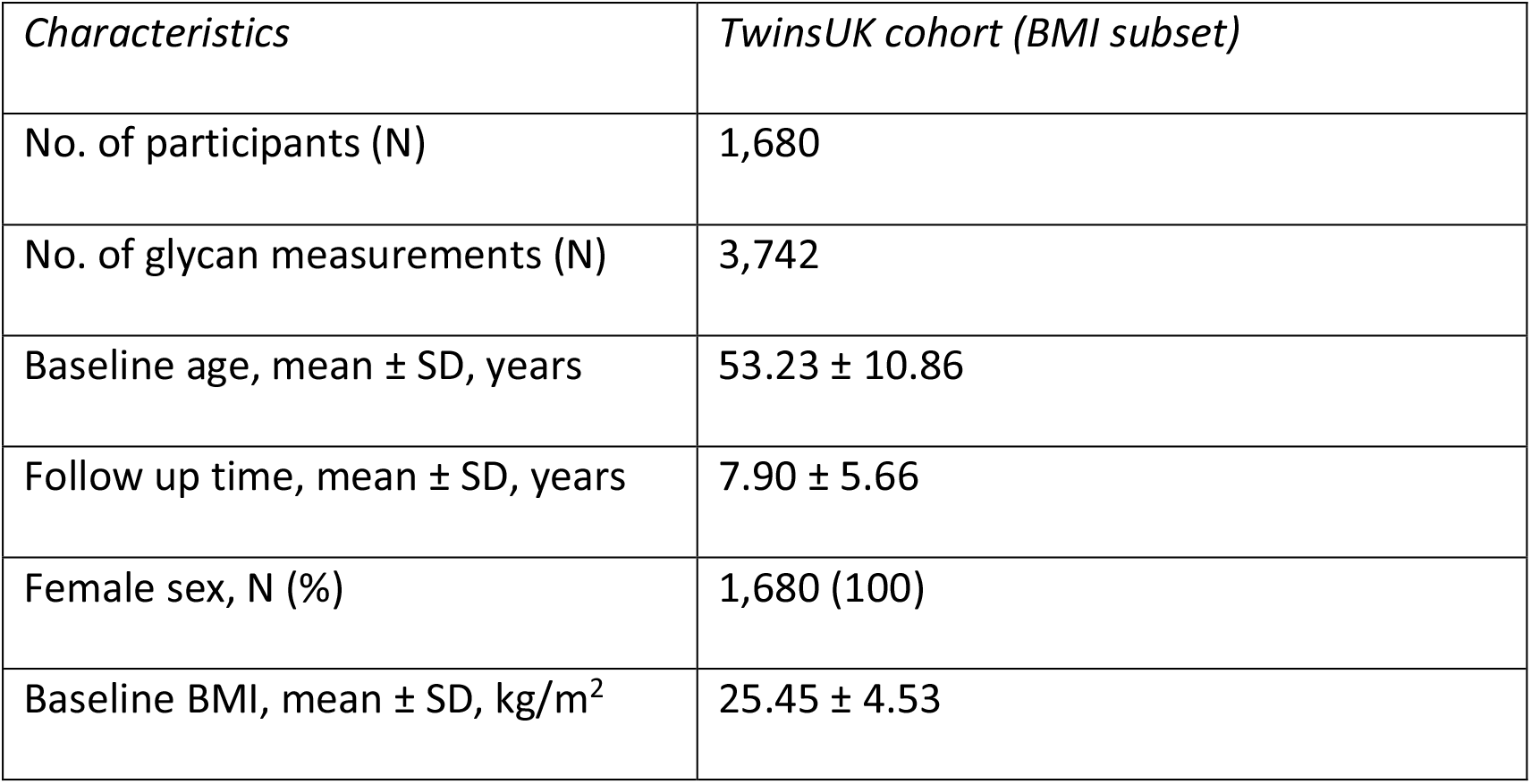
Demographic characteristics of the TwinsUK cohort.

**Table 3.** Impact of pre-surgical low-calorie diet on IgG glycosylation. Longitudinal analysis was performed by implementing a linear mixed effects model, with time as a fixed effect and the individual sample measurement as a random effect. False discovery rate was controlled using Benjamini-Hochberg method at the specified level of 0.05.

| *Derived IgG glycan trait* | *Time_effect* | *Time_SE* | *Time_p-value* | *Adjusted p-value* |
| --- | --- | --- | --- | --- |
| bisecting GlcNAc (B) | -0.2801 | 0.0743 | 0.0043 | 0.0341 |
| agalactosylation (G0) | -0.0493 | 0.1876 | 0.7930 | 0.7930 |
| monogalactosylation (G1) | 0.1122 | 0.1869 | 0.5526 | 0.7930 |
| digalactosylation (G2) | -0.0539 | 0.1387 | 0.6991 | 0.7930 |
| total sialylation (S) | 0.0941 | 0.2090 | 0.6544 | 0.7930 |
| monosialylation (S1) | 0.1354 | 0.1878 | 0.4779 | 0.7930 |
| disialylation (S2) | -0.0869 | 0.2474 | 0.7266 | 0.7930 |
| core fucosylation (CF) | 0.0810 | 0.2416 | 0.7385 | 0.7930 |

Red – significant decrease; blue – non-significant change. GlcNAc – *N*-acetylglucosamine; SE – standard error

#### IgG N-giycosylation markedly changes after weight loss surgery

Using the same chromatographic approach, we analysed samples from patients who underwent bariatric surgery. The plasma samples were collected on the day of surgery (month 0), approximately 6 months post-surgery and 12 months post-surgery. IgG N-glycans were profiled in each of these timepoints, and the obtained values were used for derived glycan traits calculations. Statistical analysis revealed extensive changes in IgG N-glycome following the bariatric procedure. Namely, four out of eight tested derived traits showed marked changes: core fucosylated (CF) and agalactosylated (G0) glycans decreased, while digalactosylated (G2) and monosialylated (S1) glycans increased after the surgery (Table 4). The IgG glycans whose abundances were increased after bariatric surgery are major components of a young IgG glycome, as they are typically associated with a younger age. The opposite applies to agalactosylated structures, which are usual denominators of an old-like IgG glycome profile. We also examined the correlation of patients’ clinical data with IgG N-glycome features using multivariate analysis, but found no statistically significant associations (Supplementary Table 4). Finally, the type of bariatric surgery (either sleeve gastrectomy or Roux-en-Y gastric bypass) did not affect IgG glycome composition. Graphical representations of the longitudinal alterations in IgG glycosylation features are depicted in Figure 1.

**Table 4.**
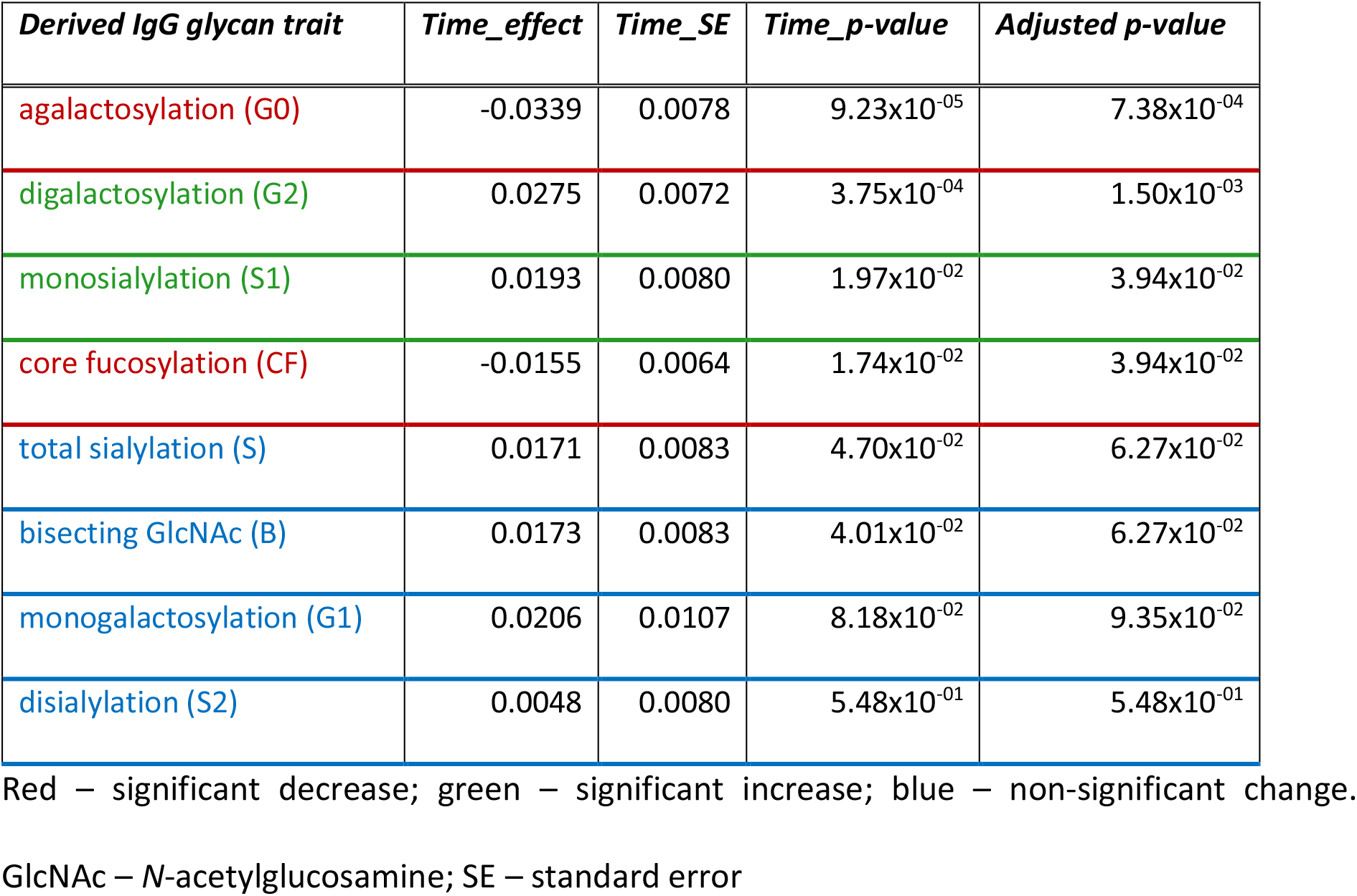
Bariatric surgery induces significant changes in IgG N-glycome. Longitudinal analysis was performed by implementing a linear mixed effects model, with time as a fixed effect and the individual sample measurement as a random effect. False discovery rate was controlled using Benjamini-Hochberg method at the specified level of 0.05.

| *Derived IgG glycan trait* | *Time_effect* | *Time_SE* | *Time_p-value* | *Adjusted p-value* |
| --- | --- | --- | --- | --- |
| agalactosylation (G0) | -0.0339 | 0.0078 | $9.23 \times 10^{-05}$ | $7.38 \times 10^{-04}$ |
| digalactosylation (G2) | 0.0275 | 0.0072 | $3.75 \times 10^{-04}$ | $1.50 \times 10^{-03}$ |
| monosialylation (S1) | 0.0193 | 0.0080 | $1.97 \times 10^{-02}$ | $3.94 \times 10^{-02}$ |
| core fucosylation (CF) | -0.0155 | 0.0064 | $1.74 \times 10^{-02}$ | $3.94 \times 10^{-02}$ |
| total sialylation (S) | 0.0171 | 0.0083 | $4.70 \times 10^{-02}$ | $6.27 \times 10^{-02}$ |
| bisecting GlcNAc (B) | 0.0173 | 0.0083 | $4.01 \times 10^{-02}$ | $6.27 \times 10^{-02}$ |
| monogalactosylation (G1) | 0.0206 | 0.0107 | $8.18 \times 10^{-02}$ | $9.35 \times 10^{-02}$ |
| disialylation (S2) | 0.0048 | 0.0080 | $5.48 \times 10^{-01}$ | $5.48 \times 10^{-01}$ |

Red – significant decrease; green – significant increase; blue – non-significant change. GlcNAc – *N*-acetylglucosamine; SE – standard error

**Figure 1.**
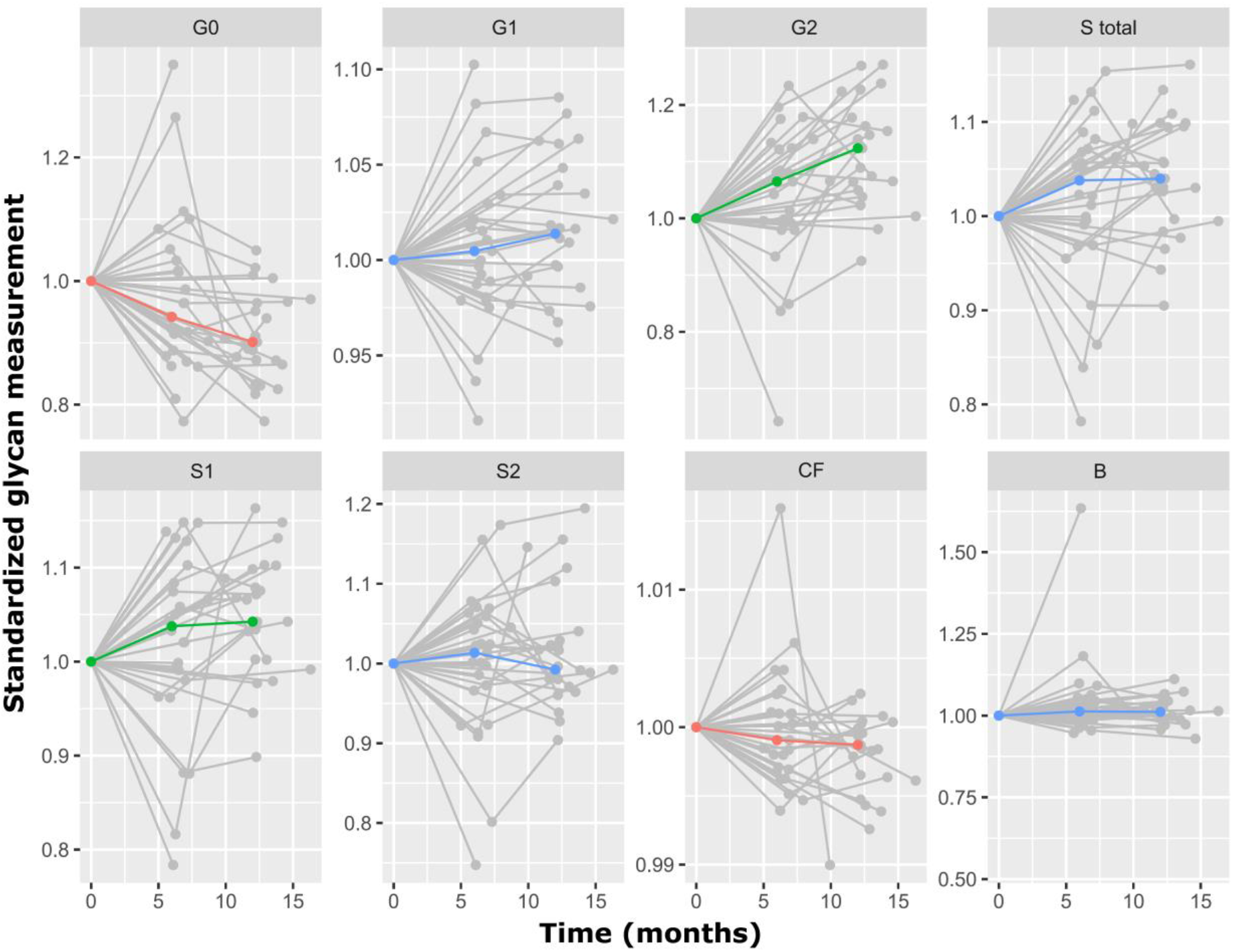
Bariatric surgery-related alterations in IgG N-glycome over time (months) Red line – significant decrease; green line – significant increase; blue line – non-significant change.

#### Weight loss induces a shift towards young-like IgG N-glycome

Using the same chromatographic approach, we profiled the plasma protein N-glycome from 1,680 TwinsUK study participants sampled at several timepoints over a 20-year-period. This served as a replication of the findings from the bariatric cohort, whose participants exhibited the reversal from old-to young-likeIgG N-glycome due to weight loss. Due to the fact that for TwinsUK cohort plasma glycome was profiled, we calculated derived traits and performed statistical analysis based only on the first 11 plasma glycan peaks, corresponding to the glycans predominantly originating from IgG [30]. We examined IgG glycome alterations associated with changes in BMI using a mixed model on a subset of 3,742 samples. Out of six examined IgG glycosylation features (derived traits), three displayed significant BMI-decrease-related changes – agalactosylation (G0), digalactosylation (G2) and incidence of high mannose structures (HM) (Table 5). Namely, the abundance of digalactosylated (G2) glycans increased with the BMI decrease, while the abundance of agalactosylated (G0) and high mannose glycans (HM) decreased with the weight loss, estimated by the BMI drop. These findings are in line with the results observed in the bariatric surgery cohort. Graphical representation of the longitudinal BMI-dependent alterations of IgG glycosylation are depicted at Figure 2.

**Table 5.** Longitudinally monitored weight loss results with significant changes of IgG N-glycosylation. Longitudinal analysis was performed by implementing a mixed model, fitted to estimate the effect of BMI change on IgG N-glycome. False discovery rate was controlled using Benjamini-Hochberg method at the specified level of 0.05.

| *Derived IgG glycan trait* | *BMI difference effect (glycan abundance (%) change per 1 kgm<sup>-2</sup> decrease in BMI)* | *BMI difference SE (glycan abundance (%) change per 1 kgm<sup>-2</sup> decrease in BMI)* | *p-value* | *Adjusted p-value* |
| --- | --- | --- | --- | --- |
| digalactosylation (G2) | 0.2004 | 0.0403 | 6.88x10<sup>-07</sup> | 5.85 x10<sup>-06</sup> |
| high mannose (HM) | -0.0519 | 0.0119 | 1.33x10<sup>-05</sup> | 5.66 x10<sup>-05</sup> |
| agalactosylation (G0) | -0.1048 | 0.0397 | 8.43x10<sup>-03</sup> | 1.79 x10<sup>-02</sup> |
| bisecting GlcNAc (B) | 0.0526 | 0.0262 | 4.49x10<sup>-02</sup> | 6.94x10<sup>-02</sup> |
| monogalactosylation (G1) | -0.0573 | 0.0343 | 9.53x10<sup>-02</sup> | 1.25x10<sup>-01</sup> |
| core fucosylation (CF) | -0.0059 | 0.0285 | 8.36x10<sup>-01</sup> | 8.89x10<sup>-01</sup> |

Red – significant decrease; green – significant increase; blue – non-significant change.
GlcNAc – N-acetylglucosamine; SE – standard error

**Figure 2.**
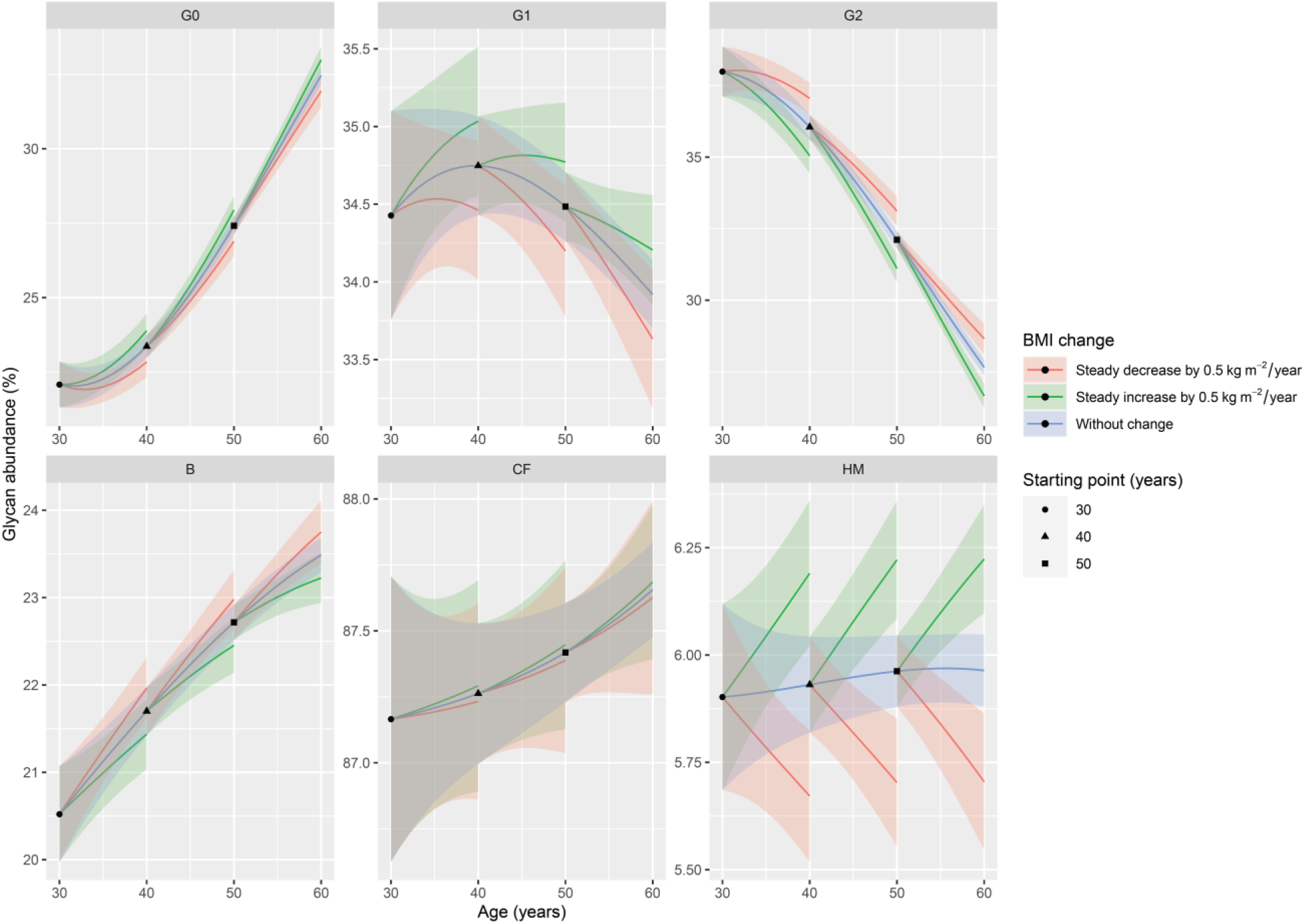
BMI-associated alterations in IgG N-glycosylation across multiple timepoints. Changes in IgG derived traits are presented with lineplots of hypothetical ageing of TwinsUK participants (all women). Black dot represents a starting point of a 30-year-old woman, black triangle of a 40-year-old woman and black square of a 50-year-old woman. All of these women have a baseline BMI of 25 kg m^-2^. Blue lines represent age-related IgG glycosylation changes attributed to stabile BMI. Green lines represent age-related IgG glycosylation changes attributed to increasing BMI (0.5 kg m^-2^ per year, through a period of 10 years). Red lines represent age-related IgG glycosylation changes attributed to decreasing BMI (0.5 kg m^-2^ per year, through a period of 10 years). Highlighted areas represent 95% confidence intervals. The effect of age on IgG glycosylation is represented with the curve slope, while the effect of BMI change is represented with the distance of green/red line from the blue line.

## DISCUSSION

In this study, we have observed extensive changes in the IgG N-glycome associated with weight loss following a low-calorie diet, bariatric surgery, or a decrease of BMI throughout time. To the best of our knowledge, this is the first study to investigate IgG N-glycome alterations in patients who underwent a low-calorie diet followed by bariatric surgery.

Prior to bariatric surgery, patients were subjected to a 3-week low-calorie diet which induced a single significant change to the IgG N-glycome. Specifically, the levels of bisecting GlcNAc were found to be reduced after the dieting period. In general, higher levels of bisecting GlcNAc are associated with enhanced affinity for FcγRs and, consequently, with enhanced antibody-dependent cellular cytotoxicity (ADCC) and other effector functions of the immune cells [9]. Hence, the reduction of bisecting GlcNAc levels on IgG decreases IgG inflammatory potential. Furthermore, several studies have reported a sex-independent increase in the levels of bisecting GlcNAc with age [32,33], suggesting that this diet-related decrease of its levels also contributes to the reduction of the biological age. Lastly, increased abundance of bisecting GlcNAc on IgG has been previously associated with type 2 diabetes [34] and with higher cardiovascular risk [13], implying that dieting improves individual’s cardiometabolic health through altered IgG glycosylation as well. As for the other examined IgG glycosylation traits, no significant changes were found to be associated with a low-calorie diet, probably due to the relatively short follow-up period (three weeks), which matches IgG serum half-life. Moreover, only 8 participants were subjected to the low-calorie diet intervention, hence, further analysis on a larger sample size might reveal additional relevant changes.

We have also analysed IgG N-glycome from individuals who underwent bariatric surgery, in a longitudinal manner. We observed several significant changes in IgG N-glycome, such as a marked decrease in agalactosylated (G0) IgG. Elevated levels of G0 IgG glycoforms are typically associated with aging, pro-inflammatory IgG glycan profile and various inflammatory diseases [9]. On the other hand, the levels of digalactosylated (G2) glycans increased after bariatric surgery and at sequential timepoints, in accordance with a reduced inflammatory potential of the circulating IgG. The increased levels of IgG galactosylation were previously associated with a younger biological age and are considered, in a way, as a measure of an individual’s well-being [9,12]. Converesely, IgG galactosylation levels substantially decrease with ageing and during inflammation [9,11]. Our results demonstrate that weight loss, resulting from bariatric surgery, can initiate the reversal from an old-like to a young-like IgG N-glycome, potentially reversing the clock for the immune/biological age. Furthermore, bariatric surgery also resulted in significant IgG glycome alteration inducing a decrease in core fucosylation. The vast majority of circulating IgG molecules bears core fucose (approximately 95%), which profoundly decreases IgG binding affinity to FcγRIIIA receptor and sequential ADCC [35]. ADCC is largely mediated by natural killer cells that can lyse target cells and fight viral infections. This would suggest that an extensive weight loss ameliorates immune responses against pathogens, by altering IgG glycosylation and modulating its effector functions. Lastly, bariatric surgery-related weight loss led to an increase in IgG sialylation, which is the main modulator of the IgG anti-inflammatory actions [36]. In addition to its anti-inflammatory actions, the level of IgG sialylation has been implicated in the pathogenesis of obesity-induced insulin resistance and hypertension, as already mentioned [20,21]. Inhibitory IgG receptor FcγRIIB was found to be expressed in the microvascular endothelium. Moreover, it was shown that hyposialylated IgG acts as its operating ligand, leading to the induction of obesity-related insulin resistance and hypertension. On the other hand, the sialylated glycoform is not activating the signalling pathways responsible for the insulin resistance and hypertension development, but is rather preserving insulin sensitivity and normal vasomotor tone, even in obese mice. Interestingly, the same group made another significant discovery – promotion of IgG sialylation breaks the link between obesity and hypertension/insulin resistance [21,22]. Namely, supplementation with the sialic acid precursor restored IgG sialylation, highlighting a potential approach to improve both metabolic and cardiovascular health in humans, with a single intervention. Our data suggest that a similar effect might be achieved by weight loss interventions, through restoring of IgG sialylation levels

In order to confirm the effects of weight loss on the biological immune age, we investigated how a decreasing BMI affects the IgG N-glycome during a 20-year-period. We observed the prominent inverse changes of agalactosylated (G0) and digalactosylated (G2) IgG glycans. Namely, agalactosylated IgG glycans significantly decreased, while digalactosylated ones substantially increased as the BMI decreased. These observations corroborated our findings from the bariatric patients, confirming that the body weight reduction reverses IgG glycome from old-like to young-like, implying at the same time a likely reduction in the biological and immune age. Nonetheless, TwinsUK participants have not experienced such an extensive weight loss, which potentially influenced the replication of other significant glycan changes from the bariatric cohort; second – herein, the weight loss was approximated by BMI decrease, which is usually a legitimate assumption, however, it does not have to apply to all cases; and third –we profiled plasma glycome, while the IgG glycan traits were approximated and the information on IgG sialylation was confounded by other plasma glycoproteins. Ideally, these issues could be circumvented in the future studies whose experimental design would allow the simultaneous, multi-centre follow-up of larger groups of patients.

Intense physical exercise can also shift IgG N-glycome towards a young-like profile by increasing IgG galactosylation [23]. Although another study reported that prolonged intensive exercise can have the opposite effect and promote pro-inflammatory changes of IgG N-glycome [37], its findings are not surprising since it recruited healthy, normal-weight female participants, subjected to the intense energy deprivation and exercise levels, to induce substantial fat loss in a rather short time period. The authors also highlighted that starting weight and the way in which weight loss is achieved could be crucial for the final effect on the immune system [37]. Therefore, it seems that exercise overall has a positive impact on the immune system and biological clock, but its intensity and duration should be personalized in order to achieve the optimal results.

To summarize, our results indicate that both dieting and bariatric surgery have an impact on inflammation and biological aging by altering IgG N-glycan patterns. All of the observed weight-loss-associated alterations in IgG glycosylation are suggesting a decreased inflammatory potential of the circulating IgG and a reduction of biological age. Hence, improving metabolic and endocrine health through weight loss consequently also contributes to the preservation of a healthy immune system.

## Data Availability

The datasets used and analysed during the current study are available from the corresponding author on reasonable request.

## ACKNOWLEDGMENTS

This research was funded by the National Institute for Health Research (NIHR) Oxford Biomedical Research Centre (BRC). The views expressed are those of the author(s) and not necessarily those of the NHS, the NIHR or the Department of Health. The authors thank Rachel Franklin, Michelle Haylock, James Chivenga, Roxanne Williams and the BRC Oxford GI Biobank for sample collection. The authors thank all the patients who took part in this study.

## AUTHORS’ CONTRIBUTION

Cristina Menni, Alessandra Geremia, Carolina V Arancibia-Cárcamo and Gordan Lauc: Conceptualization, Supervision, Project administration, Funding acquisition. Valentina L Greto, Ana Cvetko, Tamara Štambuk, Niall J Dempster, Helena Deriš, Ana Cindrić, Mario Falchi, Cristina Menni: Investigation. Domagoj Kifer, Frano Vušković: Methodology, Software, Formal analysis. Valentina L Greto, Ana Cvetko, Tamara Štambuk, Niall J Dempster, Domagoj Kifer, Frano Vučković, Mario Falchi, Jeremy W Tomlinson, Olga Gornik, Tim Spector, Cristina Menni, Alessandra Geremia, Carolina V Arancibia-Cárcamo, Gordan Lauc: Data curation and visualization: Valentina L Greto, Ana Cvetko, Tamara Štambuk, Niall J Dempster, Domagoj Kifer, Frano Vučković, Gordan Lauc: Writing-original draft. Valentina L Greto, Ana Cvetko, Tamara Štambuk, Niall J Dempster, Domagoj Kifer, Helena Deriš, Ana Cindrić, Frano Vučković, Mario Falchi, Richard S Gillies, Jeremy W Tomlinson, Olga Gornik, Bruno Sgromo, MD6, Tim D Spector, Cristina Menni, Alessandra Geremia, Carolina V Arancibia-Cárcamo, Gordan Lauc: Review & Editing.

## REFERENCES

[1] Obesity and overweight https://www.who.int/news-room/fact-sheets/detail/obesity-and-overweight (accessed April 16, 2020).

[2] Han TS, Lean ME. A clinical perspective of obesity, metabolic syndrome and cardiovascular disease. JRSM Cardiovasc Dis 2016;5:2048004016633371. https://doi.org/10.1177/2048004016633371.

[3] Alpert A, Pickman Y, Leipold M, Rosenberg-Hasson Y, Ji X, Gaujoux R, et al. A clinically meaningful metric of immune age derived from high-dimensional longitudinal monitoring. Nat Med 2019;25:487–95. https://doi.org/10.1038/s41591-019-0381-y.

[4] Franceschi C, Garagnani P, Parini P, Giuliani C, Santoro A. Inflammaging: a new immune-metabolic viewpoint for age-related diseases. Nature Reviews Endocrinology 2018;14:576–90. https://doi.org/10.1038/s41574-018-0059-4.

[5] Touch S, Clément K, André S. T Cell Populations and Functions Are Altered in Human Obesity and Type 2 Diabetes. Curr Diab Rep 2017;17:81. https://doi.org/10.1007/s11892-017-0900-5.

[6] Lauc G, Sinclair D. Biomarkers of biological age as predictors of COVID-19 disease severity. Aging 2020. https://doi.org/10.18632/aging.103052.

[7] Lauc G, Pezer M, Rudan I, Campbell H. Mechanisms of disease: The human N-glycome. Biochimica et Biophysica Acta – General Subjects 2016;1860:1574–82. https://doi.org/10.1016/j.bbagen.2015.10.016.

[8] Gornik O, Pavic T, Lauć G. Alternative glycosylation modulates function of IgG and other proteins – implications on evolution and disease. Biochim Biophys Acta 2012;1820:1318–26. https://doi.org/10.1016/j.bbagen.2011.12.004.

[9] Gudelj I, Lauc G, Pezer M. Immunoglobulin G glycosylation in aging and diseases. Cellular Immunology 2018. https://doi.org/10.1016/j.cellimm.2018.07.009.

[10] Dall’Olio F. Glycobiology of aging. Subcellular Biochemistry, 2018. https://doi.org/10.1007/978-981-13-2835-0_17.

[11] Krištić J, Vučković F, Menni C, Klarić L, Keser T, Beceheli I, et al. Glycans Are a Novel Biomarker of Chronological and Biological Ages. The Journals of Gerontology: Series A 2014;69:779–89. https://doi.org/10.1093/gerona/glt190.

[12] Štambuk J, Nakić N, Vučković F, Pučić-Baković M, Razdorov G, Trbojević-Akmačić I, et al. Global variability of the human IgG glycome. Biochemistry; 2019. https://doi.org/10.1101/535237.

[13] Menni C, Gudelj I, MacDonald-Dunlop E, Mangino M, Zierer J, Bešić E, et al. Glycosylation Profile of Immunoglobulin G Is Cross-Sectionally Associated with Cardiovascular Disease Risk Score and Subclinical Atherosclerosis in Two Independent Cohorts. Circulation Research 2018;122:1555–64. https://doi.org/10.1161/CIRCRESAHA.117.312174.

[14] Wittenbecher C, Štambuk T, Kuxhaus O, Rudman N, Vučković F, Štambuk J, et al. Plasma N-Glycans as Emerging Biomarkers of Cardiometabolic Risk: A Prospective Investigation in the EPIC-Potsdam Cohort Study. Diabetes Care 2020;43:661–8. https://doi.org/10.2337/dc19-1507.

[15] Gao Q, Dolikun M, Štambuk J, Wang H, Zhao F, Yiliham N, et al. Immunoglobulin G N-Glycans as Potential Postgenomic Biomarkers for Hypertension in the Kazakh Population. Omics 2017;21:380–9. https://doi.org/10.1089/omi.2017.0044.

[16] Liu J, Dolikun M, Štambuk J, Trbojević-Akmačić I, Zhang J, Wang H, et al. The association between subclass-specific IgG Fc N-glycosylation profiles and hypertension in the Uygur, Kazak, Kirgiz, and Tajik populations. J Hum Hypertens 2018;32:555–63. https://doi.org/10.1038/s41371-018-0071-0.

[17] Wang Y, Klarić L, Yu X, Thaqi K, Dong J, Novokmet M, et al. The Association Between Glycosylation of Immunoglobulin G and Hypertension: A Multiple Ethnic Cross-Sectional Study. Medicine 2016;95:e3379. https://doi.org/10.1097/MD.0000000000003379.

[18] Nikolac Perkovic M, Pucic Bakovic M, Kristic J, Novokmet M, Huffman JE, Vitart V, et al. The association between galactosylation of immunoglobulin G and body mass index. Progress in Neuro-Psychopharmacology and Biological Psychiatry 2014;48:20–5. https://doi.org/10.1016Zj.pnpbp.2013.08.014.

[19] Russell AC, Kepka A, Trbojević-Akmačić I, Ugrina I, Song M, Hui J, et al. Increased central adiposity is associated with pro-inflammatory immunoglobulin G N-glycans. Immunobiology 2019;224:110–5. https://doi.org/10.1016/jjmbio.2018.10.002.

[20] Sundgren NC, Vongpatanasin W, Boggan BMD, Tanigaki K, Yuhanna IS, Chambliss KL, et al. IgG receptor FcγRIIB plays a key role in obesity-induced hypertension. Hypertension 2015;65:456–62. https://doi.org/10.1161/HYPERTENSIONAHA.114.04670.

[21] Tanigaki K, Sacharidou A, Peng J, Chambliss KL, Yuhanna IS, Ghosh D, et al. Hyposialylated IgG activates endothelial IgG receptor FcγRIIB to promote obesity-induced insulin resistance. J Clin Invest 2018;128:309–22. https://doi.org/10.1172/JCI89333.

[22] Peng J, Vongpatanasin W, Sacharidou A, Kifer D, Yuhanna IS, Banerjee S, et al. Supplementation with the Sialic Acid Precursor N-acetyl-D-Mannosamine Breaks the Link Between Obesity and Hypertension. Circulation 2019. https://doi.org/10.1161/circulationaha.119.043490.

[23] Tijardović M, Marijančević D, Bok D, Kifer D, Lauc G, Gornik O, et al. Intense Physical Exercise Induces an Anti-inflammatory Change in IgG N-Glycosylation Profile. Frontiers in Physiology 2019;10:1–10. https://doi.org/10.3389/fphys.2019.01522.

[24] Nguyen NT, Kim E, Vu S, Phelan M. Ten-year Outcomes of a Prospective Randomized Trial of Laparoscopic Gastric Bypass Versus Laparoscopic Gastric Banding. Ann Surg 2018;268:106–13. https://doi.org/10.1097/SLA.0000000000002348.

[25] O’Brien P. Bariatric surgery and type 2 diabetes: a step closer to defining an optimal approach. The Lancet Diabetes & Endocrinology 2019;7:889–91. https://doi.org/10.1016/S2213-8587(19)30352-3.

[26] Verdi S, Abbasian G, Bowyer RCE, Lachance G, Yarand D, Christofidou P, et al. TwinsUK: The UK Adult Twin Registry Update. Twin Res Hum Genet 2019;22:523–9. https://doi.org/10.1017/thg.2019.65.

[27] Pavić T, Dilber D, Kifer D, Selak N, Keser T, Ljubičić Đ, et al. N-glycosylation patterns of plasma proteins and immunoglobulin G in chronic obstructive pulmonary disease. J Transl Med 2018;16. https://doi.org/10.1186/s12967-018-1695-0.

[28] Leek JT, Johnson WE, Parker HS, Jaffe AE, Storey JD. The sva package for removing batch effects and other unwanted variation in high-throughput experiments. Bioinformatics 2012;28:882–3. https://doi.org/10.1093/bioinformatics/bts034.

[29] R Core Team (2020). R: A language and environment for statistical computing. R Foundation for Statistical Computing, Vienna, Austria. URL https://www.R-project.org/. n.d.

[30] Clerc F, Reiding KR, Jansen BC, Kammeijer GSM, Bondt A, Wuhrer M. Human plasma protein N-glycosylation. Glycoconj J 2016;33:309–43. https://doi.org/10.1007/s10719-015-9626-2.

[31] Bates D, Mächler M, Bolker B, Walker S. Fitting Linear Mixed-Effects Models Using lme4. Journal of Statistical Software 2015;67:1–48. https://doi.org/10.18637/jss.v067.101.

[32] Ruhaak LR, Uh H-W, Beekman M, Koeleman CAM, Hokke CH, Westendorp RGJ, et al. Decreased Levels of Bisecting GlcNAc Glycoforms of IgG Are Associated with Human Longevity. PLOS ONE 2010;5:e12566. https://doi.org/10.1371/journal.pone.0012566.

[33] Pučić M, Knežević A, Vidič J, Adamczyk B, Novokmet M, Polašek O, et al. High Throughput Isolation and Glycosylation Analysis of IgG-Variability and Heritability of the IgG Glycome in Three Isolated Human Populations. Mol Cell Proteomics 2011;10. https://doi.org/10.1074/mcp.M111.010090.

[34] Lemmers RFH, Vilaj M, Urda D, Agakov F, Šimurina M, Klaric L, et al. IgG glycan patterns are associated with type 2 diabetes in independent European populations. Biochim Biophys Acta Gen Subj 2017;1861:2240–9. https://doi.org/10.1016Zj.bbagen.2017.06.020.

[35] Iida S, Kuni-Kamochi R, Mori K, Misaka H, Inoue M, Okazaki A, et al. Two mechanisms of the enhanced antibody-dependent cellular cytotoxicity (ADCC) efficacy of non-fucosylated therapeutic antibodies in human blood. BMC Cancer 2009;9:58. https://doi.org/10.1186/1471-2407-9-58.

[36] Kaneko Y, Nimmerjahn F, Ravetch JV. Anti-Inflammatory Activity of Immunoglobulin G Resulting from Fc Sialylation. Science 2006;313:670–3. https://doi.org/10.1126/science.1129594.

[37] Sarin HV, Gudelj I, Honkanen J, Ihalainen JK, Vuorela A, Lee JH, et al. Molecular Pathways Mediating Immunosuppression in Response to Prolonged Intensive Physical Training, Low-Energy Availability, and Intensive Weight Loss. Front Immunol 2019;10:907. https://doi.org/10.3389/fimmu.2019.00907.

